# Associations between Diabetes Self-Management and Microvascular Complications Among Patients Living in Rural Areas, in Kenya

**DOI:** 10.1101/2024.02.16.24302931

**Authors:** Rachael Ireri, Gideon Kikuvi, Susan Mambo, Besty Cheriro

## Abstract

**Introduction:** Diabetes is a major public health concern worldwide. Low and middle-income countries are the most affected. Diabetes self-management can significantly reduce the burden of diabetes complications and mortality.

**Methods:** This cross-sectional study was conducted at the outpatient department of a county referral hospital in Kenya, from 1st August 2022 to 30th October 2022. Patients with known type II diabetes of age ≥20 years visiting the hospital for routine follow-up visits were included. A 7-tem Summary of Diabetes Self-care Activities (SDSAC) Questionnaire was used to assess Diabetes self-care activities. For data entry and statistical analysis, SPSS for Windows version 27.0 was used.

**Results:** There were 96 (39.2%) males and 149 (60.8%) females. Most of the participants were more than 61 years 148 (60.4%). Prevalence of Diabetes microvascular complications among the study participants was 56.78%, 25.3%, and 14.7% for neuropathy, retinopathy, and nephropathy respectively. significant association was found between the sum scale scores of dietary activities, blood glucose testing, physical activity, foot care, and neuropathy at 95% CI and (P< 0.001). Scores on the Blood glucose testing and foot care activities subscale were significantly associated with Retinopathy (p<0.001). Additionally, significant associations were found between the presence of nephropathy and dietary activities, f00t care, and physical activity subscale (p=<0.oo1).

**Conclusions:** Diabetes self-management activities have an impact on microvascular complications in patients with diabetes.

## Introduction

Diabetes is a major public health concern that is approaching epidemic proportions worldwide(1). Levels of morbidity and mortality attributable to diabetes remain high. Though diabetes and its related complications is a global problem low and middle-income countries (LMICs) are the most affected(2). Diabetes treatment imposes a huge economic burden in developing countries and the costs are projected to rise further(3). Among the chronic complications of diabetes, microvascular complications of kidney, retina, and peripheral nerves are a significant cause of mortality and disability in patients with diabetes. This greatly affects their quality of life and care burden. the Diabetes self-management can reduce complications and mortality in patients living with diabetes(4).

Unlike previously believed that it is more of a problem of cities and towns, the rural communities have experienced an increase as well(3). The responsibility of providing preventive and therapeutic healthcare services falls on the healthcare workers in the primary health centers and dispensaries as there are not many private or tertiary-level hospitals to care for the health demands of diabetic patients(5). The situation is worse because these communities are faced with multiple challenges, including a lack of access to diabetes education, clinical services, limited smart cellphone coverage, and internet access, limited transportation and long travel distances, as well as higher rates of poverty(6).

The needs of diabetic patients are not only limited to adequate glycemic control but also prevent complications and disability(7). One of the biggest challenges for healthcare providers is addressing the continued needs and demands of individuals with diabetes(8). The regular follow-up of diabetic patients by a multidisciplinary team is vital in averting long-term complications(9,10). This is met with difficulties as more than 80% of consultants in Kenya are found in Urban areas and tertiary hospitals, this in turn contributes to suboptimal diabetes management and higher rates of diabetes-related complications(11).

### Diabetes Self-Management Activities

Self-care in diabetes is a process of development of knowledge or awareness by learning to survive with the complex nature of diabetes in a social context(12). The majority of daily diabetes care is handled by patients and/or families. Seven essential self-care behaviors in people with diabetes predict outcomes. They include; healthy eating, being physically active, monitoring of blood sugar, compliance with medications, good problem-solving skills, healthy coping skills, and risk-reduction behaviors(13–16). These measures can be useful for both clinicians and educators treating individual patients and for researchers evaluating new approaches to care(17). Self-report is an important practical and cost-effective approach for self-management assessment(18).

Diabetes self-management activities are behaviors undertaken by people with diabetes to successfully manage the disease on their own. All seven behaviors are positively correlated with good glycemic control, reduction of complications, and improvement in quality of life(9,19,20). In addition, it was observed that self-care encompasses not only performing these activities but also the interrelationships between them. Diabetes self-care requires the patient to make major lifestyle modifications enhanced with the supportive role of healthcare staff for maintaining a higher level of self-confidence leading to a successful behavior change(21–24).

Diabetes education is important but it must be transferred to action or self-care activities to fully benefit the patient. The American Association of Clinical Endocrinologists and WHO emphasize the importance of patients becoming active and knowledgeable participants in their care(10,25). Emphasis should be placed on; understanding the disease and the role of drugs, diet plans, foot, and eye care, role of continuous and sustained lifestyle modification, physical activities, and self-glucose monitoring(26).

Given the prevalence of DM and its complications in Nyeri county, there had been limited studies conducted to determine diabetes self–management among its population. Therefore, study aimed to assess the diabetes self-management practices among diabetes patients living in rural areas in Nyeri county, Kenya.

## Materials and Methods

A facility-based, cross-sectional study was conducted at the outpatient department of Nyeri County referral hospital in Kenya. The study duration was from 1st August 2022 till 30th October 2022. In this study, patients of both genders, of age 20 years and above, known cases of type II DM visiting the hospital for routine follow-up visit were included. Patients being seen in the emergency or in-patient department, patients who did not have at least one available record of HbA1c from the last six months were excluded. All patients were included after attaining informed consent. A semi-structured questionnaire was used to record patient information. It included biodata information, Duration of DM and treatment and HbA1c record from the last six months.

In order to assess their status of self-care, Summary of Diabetes self-care activities (SDSCA) Questionnaire was used. The seven items of SDSCA are used to assess self-care activities. They are divided into 7 subscales – blood glucose testing subscale (item 7,8,3A), Diet sub scale (item 1,2,3,4,1A,5A), Physical exercise subscale (item 5,6, 2A), smoking subscale (item 11,12A,13A, 14A), foot care subscale (item 9,10,9A,10A,11A), medications subscale (item6A,7A,8A) and self-care recommendations subscale (item 1A,2A,3A,4A). The questionnaire ask patient about their diabetes self-care activities during the past 7 days. If they were sick during the past 7 days, we went back to the last 7 days that they were not sick.

Scoring instructions were. Step 1: For items 1–10, use the number of days per week on a scale of 0–7. Step 2: Scoring Scales; General Diet = Mean number of days for items 1 and 2 and Specific Diet = Mean number of days for items 3, and 4 where the scale was (0=7, 1=6, 2=5, 3=4, 4=3, 5=2, 6=1,7=0); Exercise = Mean number of days for items 5 and 6; Blood-Glucose Testing = Mean number of days for items 7 and 8. Foot-Care = Mean number of days for items 9 and 10; Smoking Status = Item 11 (0 = nonsmoker,1 = smoker), and number of cigarettes smoked per day.

Scoring for Additional Items such as; Recommended regimen = Items 1A - 4A, and items 12A - 14A, no scoring required; Diet = Use total number of days for item 5A; Medications = Use item 6A - OR - 7A AND 8A, use total number of days for item 6A, use mean number of days if both 7A and 8A are applicable. Lastly; Foot-Care = Mean number of days for items 9A - 11A, after reversing 10A and including items 9 and 10.

For data entry and statistical analysis SPSS for Windows version 27 was utilized. Patient characteristics were presented as frequency and percentages and compared using Chi square test. For SDSCA overall and its subscales mean and standard deviation (SD) were calculated and for comparison independent sample t-test was applied. The p value ≤0.05 was taken as significant.

### Ethics statement

Both written and verbal informed consent for participation was also obtained from all the participants. For written consent all participants signed the consent form in the questionnaire. Verbal consent was witnessed by the nurse or clinical officer in-charge of the clinic. The study was approved by the Ethical Review Committee of Jomo Kenyatta University of Agriculture and Technology, and the National Commission for Science, Technology and Innovation (NACOSTI/P/22/18990)

## Results

### Participants characteristics, both overall and according to the presence of the microvascular complications of diabetes

Two hundred and forty-five people with Diabetes (PWD) participated in the study. Among them, 96 (39.2%) were males and 149 (60.8%) were females. Most of the participants were more than 61 years 148 (60.4%). Prevalence of Diabetes microvascular complications among the study participants were 56.78%, 25.3% and 14.7% for neuropathy, retinopathy and nephropathy respectively. Significant association was found between age and level of education with all the three microvascular complications. Furthermore, marital status was associated nephropathy and neuropathy whereas Hb1Ac was associated with nephropathy (Table 1).

**Table 1:** Participants socio-demographic characteristics association with the presence of the microvascular complications of diabetes.

|  |  |  |  |  |  |  |  |
| --- | --- | --- | --- | --- | --- | --- | --- |
| Characteristic variable | Total (n=245) | Neuropathy (n= 139; 56.7%) | P value | Retinopathy (n= 62; 25.3%) | P value | Nephropathy (n= 36 ;14.7%) | P value |
| **Age** |  |  |  |  |  |  |  |
| 20-35 | 12 (4.9%) | 1(0.7) | 0.001 | 0 (0%) | 0.009 | 2(5.6%) | 0.02 |
| 36-45 | 21 (8.6%) | 7(5.0%) |  | 3(4.8%) |  | 1(2.8%) |  |
| 46-60 | 64 (26.1%) | 27(19.4) |  | 11(17.7%) |  | 3(8.3%) |  |
| 61 and above | 148 (60.4%) | 104(74.8) |  | 48(77.4%) |  | 30(83.3%) |  |
| **Sex** |  |  |  |  |  |  |  |
| Male | 96 (39.2%) | 51 (36.7%) | 0.36 | 19 (30.6%) | 0.11 | 14 (38.9%) | 0.97 |
| Female | 149 (60.8%) | 88 (63.3%) |  | 43 (69.4%) |  | 22 (61.1%) |  |
| **Marital status** |  |  |  |  |  |  |  |
| Single | 23 (9.4%) | 7 (5.0%) | 0.003 | 3 (4.8%) | 0.08 | 2 (5.6%) | 0.007 |
| Married | 168 (68.6%) | 93 (66.9%) |  | 39 (62.9%) |  | 20 (55.6%) |  |
| Divorced/Separated | 8 (3.8%) | 4 (2.9%) |  | 2 (3.2%) |  | 0 (0%) |  |
| Widowed | 46 (18.8%) | 35 (25.2%) |  | 18 (29.0%) |  | 14 (38.9%) |  |
| **Level of Education** |  |  |  |  |  |  |  |
| Primary | 140 (57.1%) | 83 (59.7%) | 0.003 | 39 (62.9%) | 0.003 | 19 (52.8%) | 0.008 |
| Secondary | 64 (26.1%) | 28 (20.1%) |  | 7 (11.3%) |  | 6 (16.7%) |  |
| Tertiary | 13 (5.3%) | 5 (3.6%) |  | 3 (4.8%) |  | 1 (2.8%) |  |
| No schooling | 28<br>(11.4%) | 23 (16.1%) |  | 13 (21.0%) |  | 10 (27.8%) |  |
| **Occupation** |  |  |  |  |  |  |  |
| Office job | 9 (3.7%) | 3 (2.2%) | 0.17 | 3 (4.8%) | 0.13 | 0 (0%) | 0.06 |
| Outdoor job | 30<br>(12.2%) | 13 (19.4%) |  | 4 (6.5%) |  | 5 (13.9%) |  |
| Farmer | 175<br>(71.4%) | 104 (74.8%) |  | 43 (69.4%) |  | 22 (61.1%) |  |
| Stay at home | 31<br>(12.7%) | 19 (13.7%) |  | 12 (19.4%) |  | 9 (25.0%) |  |
| **Duration of diabetes** |  |  |  |  |  |  |  |
| Below 5 years | 76<br>(31.0%) | 35 (25.2%) | 0.05 | 19 (30.6%) | 0.26 | 8 (22.2%) | 0.44 |
| 5-10 years | 89<br>(36.3%) | 52 (37.4%) |  | 18 (29.1%) |  | 14 (38.9%) |  |
| More than 10 years | 80<br>(32.7%) | 52 (37.4%) |  | 25 (40.3%) |  | 14 (38.9%) |  |
| **Diabetes treatment regimen** |  |  |  |  |  |  |  |
| Oral Hypoglycemics | 165<br>(67.3%) | 96 (69.1%) | 0.73 | 41 (66.1%) | 0.90 | 21 (58.3%) | 0.27 |
| Insulin | 24<br>(9.8%) | 12 (8.6%) |  | 7 (11.3%) |  | 6 (16.7%) |  |
| Insulin plus oral hypoglycemics | 56<br>(22.9%) | 31 (22.3%) |  | 14 (22.6%) |  | 9 (25.0%) |  |
| **Hb1aC level** |  |  |  |  |  |  |  |
| Good control | 101<br>(41.2%) | 56 (40.3%) | 0.73 | 21 (33.9%) | 0.17 | 8 (22.2%) | 0.01 |
| Poor Control | 144<br>(58.8%) | 83 (59.7%) |  | 41 (66.1%) |  | 28 (77.8%) |  |
| Values are presented as number (%) and P value for associations. |  |  |  |  |  |  |  |

### Associations between Summary of diabetes self-care activities and the microvascular complications of diabetes

As shown in Table 2, a significant association was found between the sum scale scores of dietary activities, blood glucose testing, physical activity, foot care and neuropathy at 95% CI and (P< 0.001). Scores on the Blood glucose testing and foot care activities subscale were significantly associated with Retinopathy (p<0.001). Additionally, significant associations were found between the presence of nephropathy and dietary activities, f00t care and physical activity subscale (p=<0.oo1) (Table 2).

**Table: 2.**
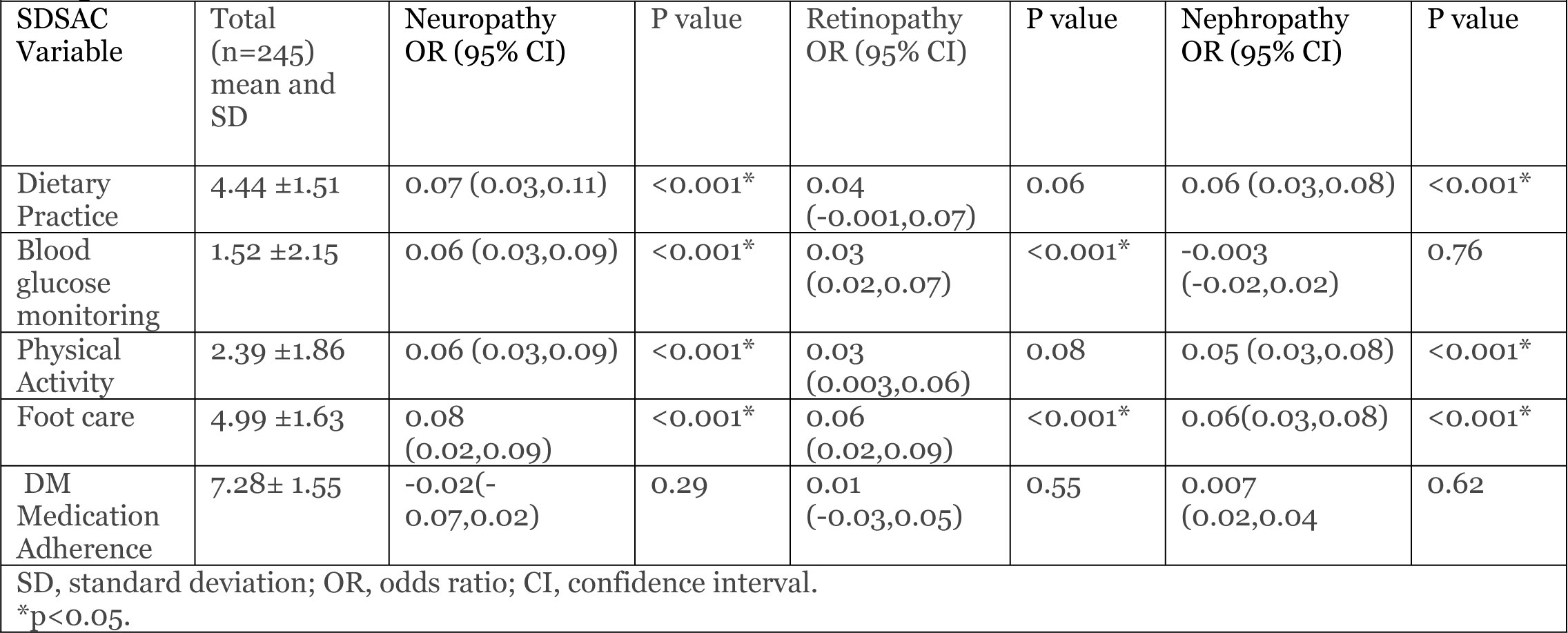
Associations between Summary of diabetes self-care activities and the microvascular plications of diabetes.

| **Table: 2 Associations between Summary of diabetes self-care activities and the microvascular complications of diabetes** |  |  |  |  |  |  |  |
| --- | --- | --- | --- | --- | --- | --- | --- |
| SDSAC Variable | Total (n=245) mean and SD | Neuropathy OR (95% CI) | P value | Retinopathy OR (95% CI) | P value | Nephropathy OR (95% CI) | P value |
| Dietary Practice | 4.44 ±1.51 | 0.07 (0.03,0.11) | <0.001* | 0.04 (-0.001,0.07) | 0.06 | 0.06 (0.03,0.08) | <0.001* |
| Blood glucose monitoring | 1.52 ±2.15 | 0.06 (0.03,0.09) | <0.001* | 0.03 (0.02,0.07) | <0.001* | -0.003 (-0.02,0.02) | 0.76 |
| Physical Activity | 2.39 ±1.86 | 0.06 (0.03,0.09) | <0.001* | 0.03 (0.003,0.06) | 0.08 | 0.05 (0.03,0.08) | <0.001* |
| Foot care | 4.99 ±1.63 | 0.08 (0.02,0.09) | <0.001* | 0.06 (0.02,0.09) | <0.001* | 0.06(0.03,0.08) | <0.001* |
| DM Medication Adherence | 7.28± 1.55 | -0.02(-0.07,0.02) | 0.29 | 0.01 (-0.03,0.05) | 0.55 | 0.007 (0.02,0.04) | 0.62 |
| SD, standard deviation; OR, odds ratio; CI, confidence interval.<br>*p<0.05. |  |  |  |  |  |  |  |

## Discussion

In this study, we sought to assess associations between diabetes self-care activities and the microvascular complications of diabetes in patients with diabetes living in rural area. In our study, Prevalence of Diabetes microvascular complications among the study participants were 56.78%, 25.3% and 14.7% for neuropathy, retinopathy and nephropathy respectively. The proportion is higher as compared to other studies(27,28). This study mainly focused on participants living in the rural area unlike other studies. This difference could be because rural populations with diabetes experience unique challenges and substantial inequities in the achievement of diabetes care(29).

In the present study, participants were further categorized into strata according to their various demographic characteristics. It was observed that patients who were older than sixty-one years were more likely to have Neuropathy, Retinopathy compared to those who were younger (p<0.05). This finding is consistent with other studies that have shown age is an important factor that affects the increased risk of prediabetes and diabetes (30). Additionally, microvascular events occur more commonly in populations with diabetes and the risk of these events are predominantly related to duration of diabetes(31).

Level of education was associated with all the MVCs. This study agrees with other studies that have revealed educational status influences the awareness about diabetes self-care, compliance to drugs and the health seeking behavior of an individual (32). Studies have observed that lower the education higher is the risk of developing diabetes complications(33).

In the present study, it was observed that dietary practices and physical activity was significantly associated neuropathy and nephropathy complications. This can be explained by a consistent healthy dietary pattern is associated with a reduced risk of microvascular complications in patients with diabetes. In this study we also assessed carbohydrate spacing and fats intake; studies have shown that high carbohydrate and high monounsaturated fat diets improve insulin sensitivity (34–36). These findings provide a reasonable basis for dietary recommendations aimed at preventing diabetic microvascular complications.

Physical activity such as walking, gardening, leisure activity, exercise, or movement for transportation or an occupation among other aerobic and resistance exercise are known to improve insulin sensitivity. Additionally, it improves the lipid profile, BMI and blood pressure. This study is consistent with other study findings that have reported that; physical activity improves glycemic control and better disease control(37–39).

Foot care practices was significantly associated with all the three microvascular complications. Others studies have demonstrated that patients with peripheral neuropathy presented with significantly higher rates of development of DR, macroalbuminuria and the existence of macrovascular complications (40–42). The results suggest that neuropathy, nephropathy and retinopathy are signs of a generalized diabetic microangiopathic process whose progression may be influenced by factors other than diabetes duration and metabolic and therefore when a patient takes measures to prevent one micro vascular complication it has direct impact to prevent of the similar complications (43,44).

The findings of association between blood glucose testing and neuropathy and retinopathy in this study are in agreement with previous literature. Blood glucose monitoring is an essential part of management in patients with diabetes mellitus (45). It helps to identify patterns in the fluctuation of blood glucose levels that occur in response to diet, exercise, medications, and pathological processes. It enhances early intervention in case of hypoglycemia or hyperglycemia which impacts on various body system such as the retina and neurons (46,47). It supports improvement of patient outcomes because critical decision can be made in time.

### Study Limitations

This study is a cross-sectional study hence cannot establish causal inferences but rather associations. Nonetheless, this study provides the extent of association between diabetes self-management and MVCs to inform development of interventions.

### Conclusion

The present study indicates that the diabetes-related self-care activities was directly linked with diabetes microvascular complications. Foot care, dietary practices, physical activity, blood glucose testing, Age, marital status, Hb1Ac and level of education were significantly associated with MVCs among patients. It is essential to promote Diabetes self-care education related to DM as a strategy to improve self-management and improve the overall patient outcome.

## Data Availability

All data is included in the tables

## Acknowledgments

Gratitude goes to workers at Diabetic Clinic and authorities of Nyeri county Referral Hospital for their great support during the study.

## Financial obligation

No funding was obtained for this research.

## Authors’ contributions

Rachael Ireri contributed to the conception of the research idea, design data analysis and interpretation, paper drafting and revision. Gideon Kikuvi contributed to the conception of the research idea, design and revision of the final draft. Susan Mambo contributed to the conception of the research idea, design, and revision of final draft. Besty Cheriro contributed to the conception of the research idea, design, and revision of final draft. All authors approved the final manuscript before publication and agree to be accountable for all aspects of the work.

## REFERENCES

1. Mathur P, Leburu S, Kulothungan V. Prevalence, Awareness, Treatment and Control of Diabetes in India From the Countrywide National NCD Monitoring Survey. Front Public Heal. 2022 Mar 14;10.

2. Sun H, Saeedi P, Karuranga S, Pinkepank M, Ogurtsova K, Duncan BB, et al. IDF Diabetes Atlas: Global, regional and country-level diabetes prevalence estimates for 2021 and projections for 2045. Diabetes Res Clin Pract. 2022 Jan 1;183.

3. Marsh Z, Teegala Y, Cotter V. Improving diabetes care of community-dwelling underserved older adults. J Am Assoc Nurse Pract. 2022 Oct 7;34(10):1156–66.

4. Tang J, Wu T, Hu X, Gao L. Self-care activities among patients with type 2 diabetes mellitus: A cross-sectional study. Int J Nurs Pract. 2021 Dec 1;27(6).

5. Alanazi M. Determinants of successful diabetes self-management behaviors among women of Arab descent with Type 2 Diabetes. Prim Care Diabetes. 2021 Apr 1;15(2):306–13.

6. Ary D V., Toobert D, Wilson W, Glasgow RE. Patient perspective on factors contributing to nonadherence to diabetes regimen. Diabetes Care. 1986;9(2):168–72.

7. Shrivastava SRBL, Shrivastava PS, Ramasamy J. Role of self-care in management of diabetes mellitus. J Diabetes Metab Disord. 2013 Mar 5;12(1).

8. Lin K, Yang X, Yin G, Lin S. Diabetes Self-Care Activities and Health-Related Quality-of-Life of individuals with Type 1 Diabetes Mellitus in Shantou, China. J Int Med Res. 2016 Feb 1;44(1):147–56.

9. Aquino JA, Baldoni NR, Flôr CR, Sanches C, Di Lorenzo Oliveira C, Alves GCS, et al. Effectiveness of individual strategies for the empowerment of patients with diabetes mellitus: A systematic review with meta-analysis. Prim Care Diabetes. 2018 Apr 1;12(2):97–110.

10. Aronson R, Brown RE, Jiandani D, Walker A, Orzech N, Mbuagbaw L. Assessment of self-management in patients with diabetes using the novel LMC Skills, Confidence and Preparedness Index (SCPI). Diabetes Res Clin Pract. 2018 Mar 1;137:128–36.

11. Afaya RA, Bam V, Lomotey AY, Afaya A. Clinical factors influencing knowledge and self-care practice among adults with type 2 diabetes mellitus. Nurs Open. 2023 Apr 1;10(4):2492–500.

12. Khosravizadeh O, Ahadinezhad B, Maleki A, Yousefy S, Momeni Z. Diabetes self-care activities among patients with type 2 diabetes: A systematic review and meta-analysis. Int J Diabetes Dev Ctries. 2023.

13. Eva JJ, Kassab YW, Neoh CF, Ming LC, Wong YY, Hameed MA, et al. Self-care and self-management among adolescent T2DM patients: A review. Front Endocrinol (Lausanne). 2018 Oct 18;9(OCT).

14. Maina PM, Pienaar M, Reid M. Self-management practices for preventing complications of type II diabetes mellitus in low and middle-income countries: A scoping review. Int J Nurs Stud Adv. 2023 Dec 1;5.

15. Young-Hyman D, De Groot M, Hill-Briggs F, Gonzalez JS, Hood K, Peyrot M. Psychosocial care for people with diabetes: A position statement of the American diabetes association. Diabetes Care. 2016;39(12):2126–40.

16. Toobert DJ, Hampson SE, Glasgow RE. The summary of diabetes self-care activities measure: Results from 7 studies and a revised scale. Diabetes Care. 2000;23(7):943–50.

17. Llera-Fábregas A, Pérez-Ríos N, Camacho-Monclova DM, Ramirez-Vick M, Andriankaja OM. Diabetes self-care activities and perception and glycemic control in adult Puerto Rican residents with Type 2 Diabetes: The LLIPDS Study. J Public health Res. 2022 Oct 1;11(4).

18. Lee J, Lee EH, Chae D, Kim CJ. Patient-reported outcome measures for diabetes self-care: A systematic review of measurement properties. Int J Nurs Stud. 2020 May 1;105.

19. Chew BH, Vos RC, Pouwer F, Rutten GEHM. The associations between diabetes distress and self-efficacy, medication adherence, self-care activities and disease control depend on the way diabetes distress is measured: Comparing the DDS-17, DDS-2 and the PAID-5. Diabetes Res Clin Pract. 2018 Aug 1;142:74–84.

20. Pintaudi B, Lucisano G, Gentile S, Bulotta A, Skovlund SE, Vespasiani G, et al. Correlates of diabetes-related distress in type 2 diabetes: Findings from the benchmarking network for clinical and humanistic outcomes in diabetes (BENCH-D) study. J Psychosom Res. 2015;79(5):348–54.

21. Kim S, Love F, Quistberg DA, Shea JA. Association of health literacy with self-management behavior in patients with diabetes. Diabetes Care. 2004 Dec;27(12):2980–2.

22. Bloomgarden Z, Gouller A. Prevention of Microvascular Complications of Diabetes. Princ Diabetes Mellit. 2004;619–38.

23. Salmerón J, Hu FB, Manson JE, Stampfer MJ, Colditz GA, Rimm EB, et al. Dietary fat intake and risk of type 2 diabetes in women. Am J Clin Nutr. 2001;73(6):1019–26.

24. Marshall JA, Bessesen DH. Dietary fat and the development of type 2 diabetes. Diabetes Care. 2002;25(3):620–2.

25. Glasgow RE, Osteen VL. Evaluating diabetes education: Are we measuring the most important outcomes? Diabetes Care. 1992;15(10):1423–32.

26. Glasgow RE, Fisher EB, Anderson BJ, LaGreca A, Marrero D, Johnson SB, et al. Behavioral science in diabetes: Contributions and opportunities. Diabetes Care. 1999 May;22(5):832–43.

27. Govindarajan Venguidesvarane A, Jasmine A, Varadarajan S, Shriraam V, Muthuthandavan AR, Durai V, et al. Prevalence of Vascular Complications Among Type 2 Diabetic Patients in a Rural Health Center in South India. J Prim Care Community Heal [Internet]. 2020 [cited 2024 Jan 29];11. Available from: 10.1177/2150132720959962

28. Pelluri R, Srikanth K, Chimakurthy J, Nagasubramanian VR. Microvascular Complications and Their Associated Risk Factors Among Rural Type 2 Diabetic Population: A Cross-Sectional Study. SN Compr Clin Med 2021 32 [Internet]. 2021 Feb 5 [cited 2024 Jan 29];3(2):625–31. Available from: https://link.springer.com/article/10.1007/s42399-021-00786-7

29. Flood D, Geldsetzer P, Agoudavi K, Aryal KK, Campos L, Brant C, et al. Rural-Urban Differences in Diabetes Care and Control in 42 Low-and Middle-Income Countries: A Cross-sectional Study of Nationally Representative Individual-Level Data. Diabetes Care [Internet]. 2022 [cited 2024 Jan 29];45. Available from: 10.2337/dc21-2342

30. Yan Z, Cai M, Han X, Chen Q, Lu H. The Interaction Between Age and Risk Factors for Diabetes and Prediabetes: A Community-Based Cross-Sectional Study. Diabetes, Metab Syndr Obes [Internet]. 2023 [cited 2024 Jan 30];16:85–93. Available from: https://www.tandfonline.com/action/journalInformation?journalCode=dmso20

31. Singh Jaggi A, Parkash Singh V, Bali A, Singh N. Advanced Glycation End Products and Diabetic Complications. Korean J Physiol Pharmacol [Internet]. 2014 [cited 2024 Jan 30];18:1–14. Available from: 10.4196/kjpp.2014.18.1.1

32. Chawla SPS, Kaur S, Bharti A, Garg R, Kaur M, Soin D, et al. Impact of health education on knowledge, attitude, practices and glycemic control in type 2 diabetes mellitus. J Fam Med Prim Care [Internet]. 2019 [cited 2024 Jan 30];8(1):261. Available from: /pmc/articles/PMC6396605/

33. Hwang Y, Lee D, Kim YS. Educational Needs Associated with the Level of Complication and Comparative Risk Perceptions in People with Type 2 Diabetes. Osong Public Heal Res Perspect [Internet]. 2020 Aug 1 [cited 2024 Jan 30];11(4):170. Available from: /pmc/articles/PMC7442446/

34. Ghaemi F, Firouzabadi FD, Moosaie F, Shadnoush M, Poopak A, Kermanchi J, et al. Effects of a Mediterranean diet on the development of diabetic complications: A longitudinal study from the nationwide diabetes report of the National Program for Prevention and Control of Diabetes (NPPCD 2016-2020). Maturitas. 2021 Nov 1;153:61–7.

35. Rydall AC, Rodin GM, Olmsted MP, Devenyi RG, Daneman D. Disordered Eating Behavior and Microvascular Complications in Young Women with Insulin-Dependent Diabetes Mellitus. N Engl J Med. 1997 Jun 26;336(26):1849–54.

36. Chudasama Y V., Khunti K. Healthy lifestyle choices and microvascular complications: New insights into diabetes management. PLoS Med. 2023 Jan 1;20(1).

37. Zhu X, Zhang X, Zhou C, Li B, Huang Y, Li C, et al. Walking pace and microvascular complications among individuals with type 2 diabetes: A cohort study from the UK Biobank. Scand J Med Sci Sport. 2023 Jan 1.

38. Makura CBT, Nirantharakumar K, Girling AJ, Saravanan P, Narendran P. Effects of physical activity on the development and progression of microvascular complications in type 1 diabetes: Retrospective analysis of the DCCT study. BMC Endocr Disord. 2013 Oct 2;13.

39. Blomster JI, Chow CK, Zoungas S, Woodward M, Patel A, Poulter NR, et al. The influence of physical activity on vascular complications and mortality in patients with type 2 diabetes mellitus. Diabetes, Obes Metab. 2013;15(11):1008–12.

40. Vithian K, Hurel S. Microvascular complications: pathophysiology and management. Clin Med (Northfield Il) [Internet]. 2010 [cited 2024 Jan 30];10(5):505. Available from: /pmc/articles/PMC4952418/

41. Mansour A, Mousa M, Abdelmannan D, Tay G, Hassoun A, Alsafar H. Microvascular and macrovascular complications of type 2 diabetes mellitus: Exome wide association analyses. Front Endocrinol (Lausanne). 2023;14.

42. Tuomilehto J, Schwarz P, Lindström J. Long-term benefits from lifestyle interventions for type 2 diabetes prevention: Time to expand the efforts. Diabetes Care. 2011 May;34(SUPPL. 2).

43. Geng T, Zhu K, Lu Q, Wan Z, Chen X, Liu L, et al. Healthy lifestyle behaviors, mediating biomarkers, and risk of microvascular complications among individuals with type 2 diabetes: A cohort study. PLoS Med. 2023 Jan 1;20(1).

44. Zhang Y, Pan XF, Chen J, Xia L, Cao A, Zhang Y, et al. Combined lifestyle factors and risk of incident type 2 diabetes and prognosis among individuals with type 2 diabetes: a systematic review and meta-analysis of prospective cohort studies. Diabetologia. 2020 Jan 1;63(1):21–33.

45. Turner R. Intensive blood-glucose control with sulphonylureas or insulin compared with conventional treatment and risk of complications in patients with type 2 diabetes (UKPDS 33). Lancet. 1998 Sep 12;352(9131):837–53.

46. Dahl-Jørgensen K, Brinchmann-Hansen O, Hanssen KF, Sandvik L, Aagenæs. Rapid tightening of blood glucose control leads to transient deterioration of retinopathy in insulin dependent diabetes mellitus: The Oslo study. Br Med J (Clin Res Ed). 1985 Mar 16;290(6471):811–5.

47. Stratton IM, Adler AI, Neil HAW, Matthews DR, Manley SE, Cull CA, et al. Association of glycaemia with macrovascular and microvascular complications of type 2 diabetes (UKPDS 35): prospective observational study. BMJ Br Med J [Internet]. 2000 Aug 8 [cited 2024 Jan 30];321(7258):405. Available from: /pmc/articles/PMC27454/

